# The efficacy and safety of hydroxychloroquine in COVID19 patients : a multicenter national retrospective cohort

**DOI:** 10.1101/2020.11.25.20234914

**Authors:** Abdulkarim Abdulrahman, Islam AlSayed, Marwa AlMadhi, Jumana AlArayed, Sara Jaafar Mohammed, Aesha Khalid Sharif, Khadija Alansari, Abdulla I AlAwadhi, Manaf AlQahtani

## Abstract

**Background:** Hydroxychloroquine is an antimalarial drug that received worldwide news and media attention in the treatment of COVID-19 patients. This drug was used based on its antimicrobial and antiviral properties despite lack of definite evidence of clinical efficacy. In this study, we aim to assess the efficacy and safety of using Hydroxychloroquine in treatment of COVID-19 patients who are admitted in acute care hospitals in Bahrain.

**Methodology:** We conducted retrospective cohort study on a random sample of admitted COVID19 patients between 24 February and 31 July 2020. The study was conducted in four acute care COVID19 hospitals in Bahrain. Data was extracted from the medical records. The primary endpoint was the requirement of non-invasive ventilation, intubation or death. Secondary endpoint was length of hospitalization for survivors. Three methods of analysis were used to control for confounding factors: logistic multivariate regression, propensity score adjusted regression and matched propensity score analysis.

**Results:** A random sample of 1571 patients were included, 440 of which received HCQ (treatment group) and 1131 did not receive it (control group). Our results showed that HCQ did not have a significant effect on primary outcomes due to COVID-19 infection when compared to controls after adjusting for confounders (OR 1.43 95% CI 0.85 to 2.37, P value=0.17). Co-administration of azithromycin had no effect on primary outcomes (OR 2.7 95% CI 0.82 to 8.85 P value =0.10). HCQ was found to be associated with increased risk of hypoglycemia (OR 10.9 95% CI 1.72 - 69.49, P value =0.011) and diarrhea(OR 2.8, 95% CI 1.4-5.5, P value =0.003), but not QT prolongation(OR=1.92, 95% CI 0.95-3.9, P value =0.06) or cardiac arrhythmia.(OR=1.06, 95% CI 0.55-2.05, P value =0.85).

**Conclusion:** Our results showed no significant beneficial effect of using hydroxychloroquine on the outcome of COVID-19 patients. Moreover, the risk of hypoglycemia due to hydroxychloroquine would possess a significant risk for out of hospital use.

## BACKGROUND AND SIGNIFICANCE

An outbreak of severe acute respiratory syndrome coronavirus-2 (SARS-CoV-2), causing the coronavirus disease 2019 (COVID19) started in December 2019, and almost a year later, we seem to be at the brink of an imminent second wave. Since it was declared a pandemic by the World Health Organisation (WHO) in March 2020 (1), it infected more than 52 million people and lead to the death of 1.3 million others (2). With no cure or vaccine identified yet, the health sector moved to repurposing available drugs.

One of the first and most rapidly identified was Hydroxychloroquine (HCQ), which was considered due to its antiviral activity. It was initially developed as an antimalarial drug, and is currently widely used to treat autoimmune diseases like systemic lupus erythematosus and rheumatoid arthritis (3). The efficacy of HCQ against SARS-CoV-2 was first confirmed *in vitro* and was reported to mediate its inhibition through the blockage of angiotensin-converting enzyme (ACE) II receptors which facilitate SARS-CoV-2 entry into cells (4). In addition, HCQ reportedly also disrupted the transport of SARS-CoV-2 from endosomes to endolysosomes, which is necessary for viral release (4, 5). HCQ also has immunomodulatory effects such as inhibition of antigen-presenting cell activity, in turn blocking the activation of T cells (6). This prevents the release of inflammatory cytokines, which causes the “cytokine storm” observed in COVID19 patients (6-8). The Food and Drug Administration issued an “emergency use authorization” for the use of HCQ for COVID19 patients, based on these limited results (9), which lead to an increase in HCQ use. The first clinical trial studying the use of HCQ to treat COVID19 was an open-label, non-randomized trial conducted in France. A total of 36 patients received HCQ and 16 controls, with results showing a drop in viral load amongst the HCQ group compared to the controls by day 6 of the trial (10). Observational studies that followed failed to report a therapeutic advantage of the magnitude seen in the French study, instead showing that HCQ has no effect on intubation or mortality amongst COVID19 patients (11, 12).

Soon after, studies showing adverse effects of HCQ use started appearing. Concerns regarding safety and efficacy increased after the infamous, and not retracted, study was published in the Lancet claiming patients treated with HCQ were at a greater risk of dying at the hospital (13). A retroscpective cohort study of 1438 patients hospitalized in metropolitan New York published in JAMA showed that patients who received HCQ (along with azithromycin) were at increased risk of cardiac arrest (11). The WHO discontinued the SOLIDARITY trial for HCQ after recommentation from the trail steering committee, based on evidence that HCQ produced little or no reduction in the mortality of hospitalized COVID-19 patients when compared to standard care (14). Results from the Randomized Evaluation of COVID-19 Therapy (RECOVERY) trial, showed that HCQ was not effective in reducing mortality and increasing length of hospital stay (15).

Results from HCQ trials and observational studies have yielded inconsistent results, making the confirmation of its efficacy difficult. This inevitably lead to a widespread confusion within the medical community and patients, with some halting its use and others continuing its administration regardless.

An increasing number of studies also reported enhanced HCQ activity when coupled with other drugs. Azithromycin, a macrolide antibiotic commonly used to treat chest infections, was reported to accelerate virus elimination (10, 16, 17). It was also used in the first HCQ clinical trial on 6 patients who, by day 6, tested negative (10). However, this was a very small sample size, and one of the patients tested positive again on day 8. The results regarding the combinations of drugs have also been inconsistent and there is no definitive proof of efficacy.

Although HCQ has a better safety clinical profile compared to chloroquine (18), the drug it is derived from, there are many reported risks and side effects of HCQ usage. Along with the common side effects, including nausea and headaches, the most common side effect of HCQ use is QT interval prolongation and subsequent risk of arrhythmia (19, 20). The mechanism by which HCQ initiates arrhythmias is yet unknown, however its electrophysiological effects include blocking several currents – funny current, L-type calcium current and rectifier potassium currents (21). These lead to sinus bradycardia and repolarization abnormalities, the later leading to the observed QT prolongation (20). A clinical trial studying the effects of different chloroquine doses involving 81 COVID19 patients in Brazil was prematurely stopped after patients receiving the higher dose (600mg, twice daily) developed arrhythmia within 2-3 days of starting the trial (22). Although chloroquine is known to be more toxic than HCQ, the study suggested that both drugs HCQ has also been associated with liver and renal impairment (8), both of which have also been reported in COVID19 patients (23). With suggestions that hepatic malfunctioning incidences increase with COVID-19 infection (24), this side effect of HCQ use could be detrimental. This, and the lack of conclusive evidence for the efficacy of HCQ in treating COVID19, creates a reluctance amongst the public and the healthcare sector to using it. This is a retrospective observational study that aims to investigate HCQ efficacy on clinical and safety outcomes amongst COVID19 patients

## METHODS

### Study design and setting

A retrospective cohort study was done on COVID19 patients in Bahrain. Cases that were admitted at Ministry of Health COVID19 treatment facilities were included. The 4 hospitals included were: Ebrahim bin Khalil Kanoo COVID19 Centre, SMC 6th floor COVID19 Centre, Hereditary Blood Disorder Centre (HBDC) COVID19 Centre and Jidhafs COVID19 Centre. All cases who were admitted to these facilities were confirmed to be infected by SARS-CoV-2 by a polymerase chain reaction (PCR) test of a nasopharyngeal sample. Cases admitted between 24 February to 31 July were included.

A random sample of cases who received HCQ and a random sample of cases who did not receive HCQ within the study time period were included. Patients who were started on NIV, intubated, died, or transferred to a different facility within 24 hours from admission were excluded from the analysis.

### Hydroxychloroquine exposure

Labeling patients as “receiving HCQ” depended on whether they were received the drug at our study baseline - defined as within 72hrs of admission. The National Bahrain treatment protocol, developed by the national task force medical team, was issued to all COVID19 facilities as guidance to health care workers for the management of COVID19. HCQ was suggested for patients with COVID19 as a therapeutic option. The suggested HCQ regimen was a loading dose of 600 mg twice on day 1, followed by 400 mg daily for 4 additional days. Azithromycin at a dose of 500 mg on day 1 and then 250 mg daily for 4 more days in combination with HCQ was an additional suggested therapeutic option. However, the suggestion of HCQ and/or Azithromycin was removed in April after several manuscripts showed lack of benefit from HCQ and a potential risk. Prescribing either or both medications was a decision left to the judgment of the treating team based on individualization of the patient care.

### Data sources and variables assessed

We obtained data from the “I-SEHA” electronic medical records. The I-SEHA is a doctor station which provides access to patient records and has all the clinical details of the hospital stay as text files. Data was manually extracted from the electronic records. 5 physicians who were assisted by 10 senior medical students reviewed all the cases and filled in an electronic form developed to collect data for this study. The data gathered included patients’ demographic details, vital signs, laboratory test results, medication lists, past medical history, clinical severity scale (as seen in the supplementary table attached in the appendix), oxygenation requirement on admission, the ratio of the oxygen saturation to the fraction of inspired oxygen (SpO2:FiO2) at admission, requirement of ICU care, ventilator use and outcomes. A complete list of variables collected is attached in Appendix A.

### Outcomes

<u>Primary outcome:</u> The primary end point was the requirement of non-invasive ventilation, intubation or death. When a patient died after ventilator requirement, the timing of the primary end point was defined as the time of the first use of ventilator.

<u>Safety outcomes:</u> the development of any of the adverse events during hospital stay, after the prescription of medications. Adverse event included were cardiac arrythmia, QT prolongation (>500ms), diarrhea and hypoglycemia (defined as glucose levels less than 3.6 mmol/L)

<u>Secondary outcome</u> was length of stay in days for survivors.

### Statistical analysis

The distribution of treatment groups was summarized. Bivariate associations between the treatment group and the measured patient characteristics were analyzed using Chi-squared (χ2) tests for categorical variables and t-test for continuous variables. We also assessed endpoint and adverse events and their associations with the treatment group.

Logistic regression model was used to estimate the relationship between HCQ use and the composite end point. A primary multivariable logistic regression model involved demographic factors, clinical factors and medications.

Propensity-score methods were used as well to reduce the effects of confounding and to account for the non-randomized treatment administration of HCQ. The individual propensities for receipt of HCQ treatment were estimated with the use of a multivariable logistic-regression model that included pre-treatment variables and predictors and risk for the outcome. Variables used were demographic factors, clinical factors and chronic diseases status.

An estimation of the association between HCQ use and the primary outcome were assessed by a multivariable logistic regression models and the use of two propensity-score methods: Propensity-score matching & the use of the propensity score as an additional covariate in the multivariate logistic regression model for the outcome.

Effect modification was examined for the primary outcome for two variables; (1) HCQ and the baseline severity of disease (whether or not patient was hypoxic), and (2) The co-prescription of azithromycin.

Estimation of the safety and secondary outcome were conducted through the use of the primary analysis, using multivariate regression models.

The STATA software, version 15.1, was used to execute the statistical analyses, (StataCorp. 2017. Stata Statistical Software: Release 15. College Station, TX: StataCorp LLC.).

### Ethical approval

The protocol and manuscript for this study were reviewed and approved by the National COVID-19 Research Committee in Bahrain (Approval Code: CRT-COVID2020-061). The National COVID-19 Research and Ethics Committee has been jointly established by the Ministry of Health and Bahrain Defence Force Hospital research committees in response to the pandemic, to facilitate and monitor COVID-19 research in Bahrain. All methods and retrospective analysis of data was approved by the National COVID-19 Research and Ethics Committee, and carried out in accordance with the local guideline and ethical guidelines of the Declaration of Helsinki 1975. All data used in this study was collected as part of normal medical procedures. Informed consent was waived by the National COVID-19 Research and Ethics Committee for this study due to its retrospective and observational nature and the absence of any patient identifying information.

## RESULTS

### Sample Characteristics

A total of 1849 cases were reviewed. Of those, 278 were excluded; 57 due to duplicates, 79 due to age less than 18 years, and 34 cases were excluded due to insufficient information. A further 56 patients were excluded due to endpoint (of ventilatory support or death) being achieved within 1 day, 7 were excluded due to transfer/discharge within 1 day, and 45 were excluded as they received HCQ out of study baseline. 1571 cases were included in the study.

Out of 1571 patients affected with COVID-19 selected in this study, 440 patients received HCQ and 1131 patients did not.

Among the patients who received HCQ. The median time to start HCQ was 1 day from admission (IQR 0-2).

Patients’ baseline characteristics (demographic and clinical) according to HCQ exposure is shown in Table 1 and 2. A propensity matched analysis was conducted to balance the two groups and their characteristics are also shown in Table 1 and 2.

**Table 1:** Unmatched and matched patient characteristics

| Factor | Level | Unmatched |  |  | Matched |  |  |
| --- | --- | --- | --- | --- | --- | --- | --- |
|  |  | Control | HCQ | p-value | Control | HCQ | p-value |
| N |  | 1131 | 440 |  | 223 | 223 |  |
| Age:, mean (SD) |  | 44.6 (15.0) | 53.4 (14.1) | <0.001 | 52.7 (14.4) | 50.5 (13.8) | 0.096 |
| male |  | 688 (60.8%) | 245 (55.7%) | 0.062 | 131 (58.7%) | 122 (54.7%) | 0.39 |
| Bahraini |  | 579 (51.2%) | 290 (65.9%) | <0.001 | 107 (48.0%) | 152 (68.2%) | <0.001 |
| Number of comorbidities | 0 | 100 (8.8%) | 40 (9.1%) | <0.001 | 17 (7.6%) | 25 (11.2%) | 0.008 |
|  | 1 | 41 (3.6%) | 43 (9.8%) |  | 10 (4.5%) | 25 (11.2%) |  |
|  | 2 | 17 (1.5%) | 27 (6.1%) |  | 7 (3.1%) | 15 (6.7%) |  |
|  | 3 | 522 (46.2%) | 135 (30.7%) |  | 83 (37.2%) | 66 (29.6%) |  |
|  | 4+ | 451 (39.9%) | 195 (44.3%) |  | 106 (47.5%) | 92 (41.3%) |  |
| Sickle Cell Disease |  | 26 (2.3%) | 6 (1.4%) | 0.24 | 2 (0.9%) | 5 (2.2%) | 0.25 |
| G6PD Deficiency |  | 134 (11.8%) | 29 (6.6%) | 0.002 | 18 (8.1%) | 22 (9.9%) | 0.51 |
| Diabetes Mellitus |  | 273 (24.1%) | 174 (39.5%) | <0.001 | 72 (32.3%) | 74 (33.2%) | 0.84 |
| Cardiovascular Disease (CVD) |  | 105 (9.3%) | 53 (12.0%) | 0.10 | 23 (10.3%) | 27 (12.1%) | 0.55 |
| Hypertension |  | 285 (25.2%) | 176 (40.0%) | <0.001 | 79 (35.4%) | 78 (35.0%) | 0.92 |
| Asthma |  | 42 (3.7%) | 23 (5.2%) | 0.18 | 9 (4.0%) | 12 (5.4%) | 0.50 |
| Chronic Obstructive Pulmonary Disease (COPD) |  | 2 (0.2%) | 4 (0.9%) | 0.035 | 0 | 0 |  |
| Obesity (BMI >=30) |  | 33 (2.9%) | 20 (4.5%) | 0.11 | 7 (3.1%) | 13 (5.8%) | 0.17 |
| Chronic Kidney Disease (CKD) |  | 37 (3.3%) | 22 (5.0%) | 0.11 | 12 (5.4%) | 13 (5.8%) | 0.84 |
| Other Chronic Lung Disease (Not asthma nor COPD) |  | 4 (0.4%) | 2 (0.5%) | 0.77 | 2 (0.9%) | 1 (0.4%) | 0.56 |
| Smoker | Current | 18 (1.9%) | 11 (2.6%) | 0.52 | 3 (1.6%) | 7 (3.3%) | 0.30 |
|  | Exsmoker | 14 (1.5%) | 4 (0.9%) |  | 3 (1.6%) | 1 (0.5%) |  |
|  | Never | 921 (96.6%) | 414 (96.5%) |  | 182 (96.8%) | 207 (96.3%) |  |

**Table 2:** Unmatched and matched patient clinical characteristics

|  |  | Unmatched |  |  | Matched |  |  |
| --- | --- | --- | --- | --- | --- | --- | --- |
| Factor | Level | Control | HCQ | p-value | Control | HCQ | p-value |
| N |  | 1131 | 440 |  | 223 | 223 |  |
| Symptoms on admission: |  | 699 (61.8%) | 303 (68.9%) | 0.009 | 154 (69.1%) | 151 (67.7%) | 0.76 |
| Fever (>38C) |  | 230 (20.3%) | 118 (26.8%) | 0.005 | 48 (21.5%) | 49 (22.0%) | 0.91 |
| Cough |  | 450 (39.8%) | 218 (49.5%) | <0.001 | 108 (48.4%) | 107 (48.0%) | 0.92 |
| Chest Pain |  | 91 (8.0%) | 49 (11.1%) | 0.054 | 23 (10.3%) | 26 (11.7%) | 0.65 |
| Shortness of Breath |  | 210 (18.6%) | 85 (19.3%) | 0.73 | 53 (23.8%) | 48 (21.5%) | 0.57 |
| Loss of smell |  | 30 (2.7%) | 18 (4.1%) | 0.14 | 4 (1.8%) | 14 (6.3%) | 0.016 |
| Loss of taste |  | 30 (2.7%) | 17 (3.9%) | 0.21 | 6 (2.7%) | 14 (6.3%) | 0.067 |
| Diarrhea |  | 61 (5.4%) | 30 (6.8%) | 0.28 | 14 (6.3%) | 16 (7.2%) | 0.71 |
| Nausea or Vomiting |  | 46 (4.1%) | 34 (7.7%) | 0.003 | 9 (4.0%) | 17 (7.6%) | 0.11 |
| Body pain |  | 167 (14.8%) | 89 (20.2%) | 0.008 | 37 (16.6%) | 49 (22.0%) | 0.15 |
| Heart Rate on admission: bpm, mean (SD) |  | 85.8 (13.7) | 86.4 (13.6) | 0.44 | 85.4 (14.3) | 86.6 (13.0) | 0.35 |
| SBP on admission: mmHg, mean (SD) |  | 129.6 (18.4) | 133.7 (18.3) | <0.001 | 132.4 (20.7) | 133.6 (18.8) | 0.50 |
| DBP on admission: mmHg, mean (SD) |  | 77.9 (11.5) | 76.8 (11.3) | 0.080 | 78.7 (10.9) | 77.5 (11.2) | 0.25 |
| Requirment of Oxygen support on admission |  | 104 (9.2%) | 54 (12.3%) | 0.069 | 28 (12.6%) | 27 (12.1%) | 0.89 |
| Oxygenation device on admission | Nasal Canula | 43 (41.0%) | 36 (66.7%) | 0.008 | 8 (29%) | 17 (63%) | 0.032 |
|  | Face Mask | 45 (42.9%) | 14 (25.9%) |  | 16 (57%) | 7 (26%) |  |
|  | Nonrebreather Face mask | 17 (16.2%) | 4 (7.4%) |  | 4 (14%) | 3 (11%) |  |
| SpO2:FiO2 ratio, mean (SD) |  | 445.08 (71.63) | 439.82 (71.32) | 0.19 | 436.17 (82.08) | 438.35 (75.45) | 0.77 |
| Presence of an elevated ALT>40U/L on admission |  | 249 (23.6%) | 114 (26.8%) | 0.19 | 49 (23.2%) | 70 (32.0%) | 0.043 |
| Presence of an elevated Creatinine on admission |  | 78 (6.9%) | 52 (11.8%) | 0.001 | 23 (10.3%) | 25 (11.2%) | 0.76 |
| Chest Xray findings on admission | Pneumonia | 291 (31.0%) | 167 (39.5%) | 0.002 | 59 (31.4%) | 73 (33.5%) | 0.65 |
|  | Normal | 648 (69.0%) | 256 (60.5%) |  | 129 (68.6%) | 145 (66.5%) |  |
| Hypotension (SBP<90mmHg or DBP<60mmHg) on admission |  | 43 (3.8%) | 14 (3.2%) | 0.56 | 7 (3.1%) | 7 (3.1%) | 1.00 |
| Tachypnea (RR>22) on admission |  | 20 (1.8%) | 11 (2.5%) | 0.35 | 5 (2.2%) | 8 (3.6%) | 0.40 |
| Baseline clinical severity scale | Isolated and asymptomatic | 5 (0.4%) | 1 (0.2%) | <0.001 | 0 (0.0%) | 1 (0.4%) | 0.33 |
|  | Isolated Mild symptomatic | 4 (0.4%) | 1 (0.2%) |  | 2 (0.9%) | 1 (0.4%) |  |
|  | Admitted on Room Air | 1018 (90.0%) | 363 (82.5%) |  | 193 (86.5%) | 182 (81.6%) |  |
|  | Admitted and on Oxygen support | 104 (9.2%) | 75 (17.0%) |  | 28 (12.6%) | 39 (17.5%) |  |
|  | Admitted and on NIV/HFNC | 0 | 0 |  | 0 | 0 |  |
|  | Admitted and on Mechanical ventilation/ECMO | 0 | 0 |  | 0 | 0 |  |
| Azithromycin during hospital stay: |  | 150 (13.3%) | 236 (53.6%) | <0.001 | 59 (26.5%) | 64 (28.7%) | 0.60 |
| Kaletra during the hospital stay |  | 186 (16.4%) | 82 (18.6%) | 0.30 | 38 (17.0%) | 39 (17.5%) | 0.90 |
| Ribavirin during the hospital stay |  | 180 (15.9%) | 45 (10.2%) | 0.004 | 33 (14.8%) | 29 (13.0%) | 0.58 |
| Steroids during hospital stay |  | 98 (8.7%) | 66 (15.0%) | <0.001 | 19 (8.5%) | 39 (17.5%) | 0.005 |
| Tocilizumab during hospital stay |  | 31 (2.7%) | 29 (6.6%) | <0.001 | 6 (2.7%) | 15 (6.7%) | 0.044 |
| Received Convalescent Plasma Transfusion during hospital stay |  | 19 (1.7%) | 33 (7.5%) | <0.001 | 3 (1.3%) | 17 (7.6%) | 0.001 |

In the unmatched sample, patients who received HCQ had a significantly higher mean age (43.4 years), were more likely to be Bahraini and had more comorbidities. Diabetes and hypertension were more common in patients receiving HCQ. The HCQ-receiving patients were more likely to be symptomatic (68.9% compared to 61.8%). Symptoms of fever, cough, body ache, nausea and vomiting were more predominant in patients who received HCQ. The HCQ-receiving patients were also more severely ill on admission, as 12.3% received supplemental oxygen on admission (through nasal cannula, face mask and Nonrebreather mask).

### The Propensity score

The distribution of the estimated propensity scores for receiving HCQ among patients who did and did not receive HCQ is shown in Appendix B. The C-statistic of the propensity-score model was 0.83. In the matched analytic sample, 223 patients were exposed to HCQ and 223 were not exposed. The differences between HCQ and pretreatment variables were attenuated in the propensity-score–matched samples as compared with the unmatched samples.

### Primary outcome

During the period of their admission, patients who received HCQ were more likely to develop the composite outcome. 24 of 440 patients (5.45%) receiving HCQ developed the primary outcome of requiring ventilatory support (invasive and non-invasive) or death in comparison to the 44 of 1131 patients (3.89%) who were not treated with HCQ. Table 3 summarizes outcomes in each treatment group.

**Table 3:** Outcomes within the unmatched and matched samples

|  | Unmatched |  |  | Matched |  |  |
| --- | --- | --- | --- | --- | --- | --- |
| Factor | Control | HCQ | p-value | Control | HCQ | p-value |
| N | 1131 | 440 |  | 223 | 223 |  |
| Primary outcome : Ventilation or Death | 44 (3.9%) | 24 (5.5%) | 0.17 | 7 (3.1%) | 12 (5.4%) | 0.24 |
| Ventilation | 41 (3.6%) | 24 (5.5%) | 0.10 | 7 (3.1%) | 12 (5.4%) | 0.24 |
| Invasive ventilation | 18 (1.6%) | 9 (2.0%) | 0.53 | 3 (1.3%) | 6 (2.7%) | 0.31 |
| Death | 26 (2.3%) | 8 (1.8%) | 0.56 | 6 (2.7%) | 5 (2.24%) | 0.76 |
| Cardiac Arrythmia: | 38 (3.4%) | 19 (4.3%) | 0.36 | 10 (4.5%) | 9 (4.0%) | 0.81 |
| QT prolongation of more than 500msec | 33 (2.9%) | 22 (5.0%) | 0.044 | 5 (2.2%) | 13 (5.8%) | 0.054 |
| Adverse events: Hypoglycemia <3.9 / 70 | 2 (0.2%) | 7 (1.6%) | <0.001 | 1 (0.4%) | 4 (1.8%) | 0.18 |
| Adverse events: Diarrhea | 23 (2.0%) | 27 (6.1%) | <0.001 | 5 (2.2%) | 14 (6.3%) | 0.035 |

The difference between the two groups was not significant across the different methods used to control confounders. The primary analysis using multivariate model showed an odds ratio of 1.43 with a 95% CI 0.85 to 2.37, P value=0.17. Other methods of confounding adjustment showed similar and non-significant results. Table 4 summarizes the analysis results.

**Table 4:** Risks for developing the primary outcome

| **Analysis** | **Ventilation or Death** | **P Value** |
| --- | --- | --- |
| **No. of events/no. of patients at risk (%)** | 68/1571(4.3%) | - |
| **Hydroxychloroquine** | 24/440 (5.45%) | - |
| **No Hydroxychloroquine** | 44/1131(3.89%) | - |
| **Crude analysis – Odds Ratio (95%CI)** | 1.43 (0.85 – 2.37) | 0.17 |
| **Multivariable analysis – Odds Ratio (95%CI)** | 1.65 (0.81 – 3.32) | 0.16 |
| **Propensity score Analysis – Odds Ratio (95%CI)** |  |  |
| **With Matching** | 1.75 (0.68 – 4.54) | 0.24 |
| **With Adjusted for Propensity score** | 0.87 (0.47 – 1.64) | 0.67 |

There was a significant effect modification in COVID-19 patients receiving HCQ and requiring supplemental oxygen on admission. Significant effect modification was also noted on in cases exposed to azithromycin. Appendix C shows the detailed effect modification analysis.

179 patients required supplemental oxygen on admission (nasal cannula, face mask or nonrebreather face mask). 75 patients received HCQ and 104 did not. Patients who were treated by HCQ were less likely to develop the outcome if they required oxygen on baseline. 15 patients in the HCQ group developed the primary outcome (20%), compared to 30 in the control group (28.85%). The difference was non-significant in the primary analysis (OR 1.09 95% CI 0.38 – 3.07).

1392 patients were admitted on room air and did not require supplemental oxygen. Of those, 365 received HCQ and 1027 did not. 9 patient who received HCQ developed the primary outcome (2.47%), while 14 patients in the control group developed the outcome (1.36%). Treatment with HCQ showed a non-significant increase in odds ratio to develop the primary outcome (OR 2.79 95% CI 0.92 to 8.43). Table 5 summarizes the results stratified by oxygen requirement at baseline.

**Table 5:** Risks for developing the primary outcome, in cases who required and did not require supplemental oxygen at baseline

| Analysis | Requiring Oxygen at Baseline | Not Requiring Oxygen at Baseline |
| --- | --- | --- |
| No. of events/no. of patients at risk (%) | 45/179 (25.14%) | 23/1392 (1.65%) |
| Hydroxychloroquine | 15/75 (20.00%) | 9/365 (2.47%) |
| No Hydroxychloroquine | 30/104 (28.85%) | 14/1027 (1.36%) |
| Crude analysis – Odds Ratio (95%CI) | 1.09 (0.38 – 3.07) | 2.79 (0.92 – 8.43) |

The analysis showed insignificant results when stratified by azithromycin exposure. It was noted that patients who received HCQ and azithromycin had an Odds ratio of 2.7 to develop the primary outcome (95% CI 0.82 to 8.85). Patients who were treated by HCQ and did not receive azithromycin had an Odds ratio of 1.3 to develop the primary outcome (95% CI 0.44 to 3.75). Results are summarized in Table 6.

**Table 6:**
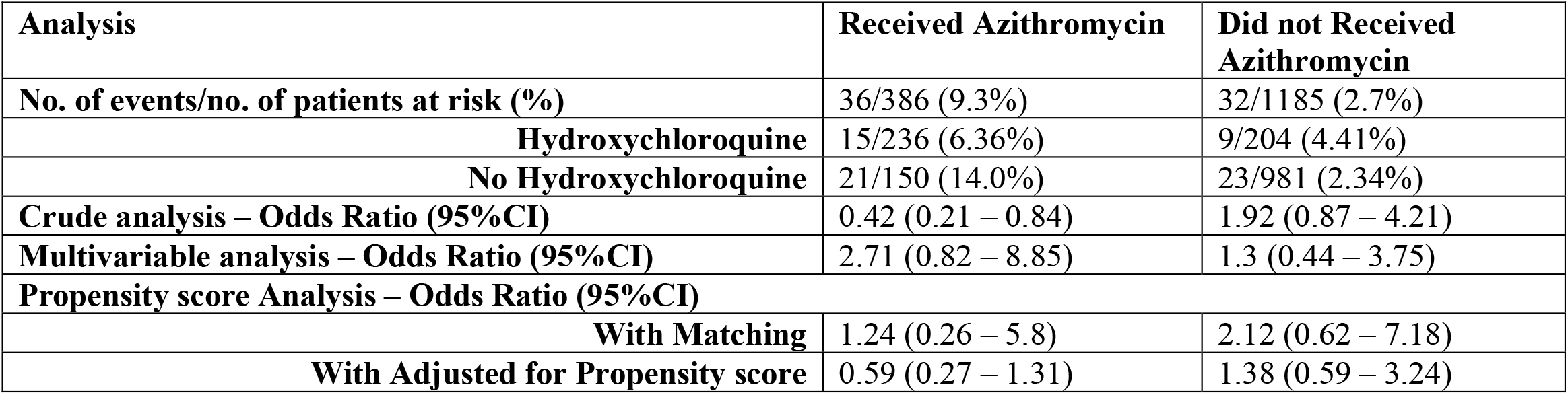
Risks for developing the primary outcome, in cases who received and did not receive azithromycin

### Safety outcome

Patients who received HCQ had significantly increased odds ratio to develop hypoglycemia (OR=10.9, 95% CI 1.72-69.49, P value =0.011) and diarrhea (OR=2.8, 95% CI 1.4-5.5, P value =0.003). Patient treated with HCQ had more patients developing QT prolongation (OR=1.92, 95% CI 0.95-3.9, P value =0.06) and cardiac arrhythmias (OR=1.06, 95% CI 0.55-2.05, P value =0.85) however these findings were non-significant. Table 7 summarizes the safety endpoints.

**Table 7:** Safety outcomes and adverse events

| Multivariable Analysis | Odds Ratio (95% CI) | P Value |
| --- | --- | --- |
| Hypoglycemia | 10.9 (1.72 – 69.49) | 0.011 |
| Diarrhoea | 2.8 (1.4 – 5.5) | 0.003 |
| QT Prolongation | 1.92 (0.95 – 3.9) | 0.06 |
| Cardiac Arrhythmia | 1.06 (0.55 – 2.05) | 0.85 |

### Secondary outcome: Length of stay in survived cases

The mean length of stay of discharged patients in the study cohort was 10.0 days (+/-5.54 days). The minimum stay was 2 days, and the maximum was 57. Patients who received HCQ had a mean stay of 11.3 days (5.65 days) while patients in the control group had mean stay of 9.5 days (5.41 days). The difference was statistically significant in a two-sided t-test (p<0.001). After adjustment for confounders using a multivariate model, HCQ had a higher length of stay by 0.63 days, however this difference was non-significant (95% CI ranged from −0.02 to 1.29). Table 8 summarizes these findings.

**Table 8:** Length of stay analysis

| Analysis | Value |  |
| --- | --- | --- |
| Overall Mean length of stay for survivors (+/- SD) | 10.0 (5.54) | - |
| Length of stay in survivor in Hydroxychloroquine group - mean in days (SD) | 11.3 (5.65) | - |
| Length of stay in survivor in the control group - mean in days (SD) | 9.5 (5.41) | - |
| Two Sample T test: Difference | - 1.77 | P<0.001 |
| Multivariable analysis – Odds Ratio (95%CI) | 0.63<br>(95% Ci : -0.02 to 1.3) | P=0.058 |

Regression models results and details are attached in Appendix C

## DISCUSSION

The results of this study show that, for our studied sample and models used, HCQ did not have a significant effect on primary outcomes (requirement for ventilation or death) due to COVID-19 infection.

Analysis of the demographics of our studied sample showed that patients who received HCQ were significantly of older age, which could be associated with a more severe HCQ-requiring presentation. Although this is an extrapolation from our data, age has reportedly been associated with a more severe progression of the disease (25-28). However, a recent study quantifying the isolated effect of age on severity of COVID-19 outcomes concluded a minimal influence of age after adjusting for important age-dependent risk factors (eg: diabetes, hypertension, cardiovascular disease (CVD) etc.) (29). Indeed, this was observed in our HCQ-receiving cohort, which showed a higher number of associated comorbidities namely diabetes mellitus, hypertension and COPD compared to the control cohort. A meta-analysis of 34 studies conducted by *Zhou et al*. showed that chronic comorbidities increase the risk of severe course and progression of the disease, with strong correlations with hypertension, diabetes and CVD (30). However, there was no difference in rates of CVD presentation between the HCQ-receiving and control groups, which may be due to increased prevalence of CVD in the country (31). The HCQ receiving group had significantly less G6PD deficiency, stemming from management guidelines contraindicating HCQ in patients with G6PD deficiency due to increased risk of hemolytic crisis (32, 33).

Patients who received HCQ had a higher presentation of symptoms on admission and scored significantly higher on baseline clinical severity scale. Creatinine levels were significantly elevated amongst patients who received HCQ, indicating COVID-19-mediated acute kidney injury (34). A significantly higher proportion of HCQ receiving patients presented with chest x-ray findings of pneumonia compared to control patients. All these presentations on admission indicate a more severe progression of the disease that is a risk of poor prognosis (28, 34, 35), increasing risk of developing composite outcome and hence indicating HCQ requirement. This may also explain the higher incidence of composite outcomes seen amongst patients receiving HCQ.

Interestingly we found no difference in requirement of oxygenation on admission between the HCQ and control groups in our studied sample. This was not expected as requirement of supplemental oxygenation on admission has been associated with increased risk of severe illness (36), and hence expected to be prescribed HCQ. However, this could be interpreted alongside the increased G6PD deficiency amongst the control groups. With a high prevalence of G6PD in Bahrain (37), it may be that many severe COVID-19 admission, that potentially required oxygen on admission, were contraindicated to receive HCQ. Yet, more patients required oxygenation on baseline in the hydroxychloroquine group. This indicates that physicians tends to prescribe HCQ in the sicker patients.

Almost all factors were insignificant after propensity score matching analysis. However, it is important to note that to conduct matched analysis, the sample size was reduced significantly. The effect of HCQ on the development of the primary outcome remained insignificant using the various ways mentioned to control for confounders.

There was no significant difference in the clinical outcome between HCQ and control groups of patients with mild to moderate disease who did not require oxygen at baseline. Therefore, there is insufficient evidence to suggest benefit of using HCQ to treat patients with low risk of developing severe disease. This finding was consistent across several reports (38, 39).

Patients in our study who received HCQ while on oxygen therapy had lower rates of developing the primary outcome, yet this as still non-significant.

Our study showed no clinical benefit from using HCQ in COVID19 patients. Moreover, the effect remained non-significant across different subgroups: room air/oxygen therapy and with and without azithromycin cotreatment.

Our study showed no benefit from combination of azithromycin with HCQ. Other studies done in France (10) and Brazil (38) also supported our outcome, and showed no clinical benefit in using the combination of HCQ and azithromycin in the treatment of COVID-19 patients.

Our study showed that HCQ does not affect the length of hospitalization. The raw analysis showed a significantly higher length of stay compared to patients not receiving HCQ, consistent with several reports (15, 40). This is explained by the more severe presentation, higher comorbidities, and risk of lower prognosis leading to the administration of HCQ. Hence, patients who received HCQ would be expected to require a longer stay. Indeed, when these factors were adjusted for in the analysis, the difference was non-significant, which is consistent with other reported data (41). These findings were also reported in a randomized clinical trial conducted in china on 150 COVID19 patients. The findings in the trial did not provide evidence to support an increase in the probability of negative conversion of SARS-CoV-2 conferred by the addition of HCQ to the standard of care in patients admitted to the hospital with COVID19.(39)

With the use of HCQ there was a significant risk of developing adverse effects, specifically hypoglycemia and diarrhea. Due to their mechanism of action, it has been well known that antimalarials cause hypoglycemia. A few studies showed the role of HCQ in diabetic patients and showed a decreased requirement in insulin (42, 43). As for diarrhea, it is a known adverse effect of HCQ as well (44). The increased risk of hypoglycemia is alarming, as it would potentially be of a significant risk if patients prone to hypoglycemia or are receiving HCQ outside hospital setting.

The surprising result was the insignificant association between QT interval prolongation and the use of HCQ. It was difficult to find a study that supported our result, as most studies showed frequent prolongation of the QT segment (45). Our result can be explained by the local protocol used in our hospitals. As daily ECG was done for all patients on HCQ. The local protocol suggests withholding HCQ once QT exceed 470ms and can then be restarted once QT has decreased. Moreover patients with a baseline QT >470msec or those who are at risk for developing cardiac arrythmia or QT prolongation are seldomly prescribed HCQ.

The findings in our study are supported by findings from multiple clinical trials and observational studies. The RECOVERY trail which randomized 4716 patients across 176 hospitals in the United Kingdom. This trial showed that HCQ had no benefit in decreasing mortality nor invasive ventilation. The findings were consistent across different subgroups includes those who received and didn’t receive oxygen at baseline(15). Another trail conducted in the US which randomized 479 patients to determine if HCQ improved clinical outcomes at 14 days also supported our results. The trail was multicentered, double blinded, placebo-controlled study. The study concluded that HCQ didn’t improve clinical outcomes in patient with COVID19 respiratory illness. These findings were consistent in all subgroups and for all outcomes evaluated, including an ordinal scale of clinical status, mortality, organ failures, duration of oxygen use, and hospital length of stay (46). A randomized clinical trial in Brazil was conducted on 667 mild-moderate COVID19 patients to measure the effect of HCQ with or without azithromycin on the clinical status at 15 days. The trial concluded that the use of HCQ, alone or with azithromycin, did not improve clinical status at 15 days as compared with standard care (38). A large observational study was conducted on 1438 hospitalized patients in COVID19 patients in New York State to measure the effect of HCQ, with or without azithromycin on the mortality rates. The study concluded that HCQ, azithromycin, or both, compared with neither treatment, was not significantly associated with differences in in-hospital mortality in COVID19 patients (11).

Interpreted along with these prior studies, the results of this study provide additional evidence that HCQ is not beneficial for adults hospitalized with COVID-19. Admitted and on Oxygen support

## STRENGTHS

The study has several strengths. It involved majority of hospitals which provide acute care for hospitalized COVID19 cases. Moreover, our study included all hospitals that use HCQ as part of the treatment regimen. The data collection process was done manually and hence all patients files were reviewed carefully and all documented details were collected. The outcomes and adverse were collected after the medication starting date, any event occurring within 24hour of admission or prior to starting the study drug were excluded.

## LIMITATIONS

The study main limitation is its design, being a retrospective observational study. Secondly, given the retrospective design, information that wasn’t documented weren’t available for analysis, and these can be potential confounders. These included : time from symptom onset, inflammatory markers. It is also likely that there is still unmeasured residual confounding due to factors not included in the analysis.

## CONCLUSION

Our results showed no significant beneficial effect of using HCQ on the outcome of COVID-19 patients. Moreover, the risk of hypoglycemia due to HCQ would possess a significant risk for out of hospital use.

## Data Availability

All the data for this study will be made available upon reasonable request to the corresponding author.

## Declarations

### Ethics approval and consent to participate

The study was approved by the National COVID-19 Research and Ethics Committee.

### Consent for publication

All authors gave their consent for publication.

### Author contributions

IS, JA, KA, SJ, AKS, MA gathered the data and supervised the data collection team. AA and AIA analyzed the data. AA, MA, JA, SJ wrote the manuscript. AA and MQ interpreted data and edited the manuscript. All authors reviewed and approved the final version of the manuscript. Manaf Alqahtani is the guarantor of this work.

### Corresponding author

Manaf AlQahtani

Phone: +973 39766000

## Acknowledgements

We would like to express our gratitude towards our colleagues: Ammar Kheyami, Mujtaba Mal Alla, Abdulla AlMuharraqi, Zeyad Mahmood, Narjis Ali AlSheala, Ola Husain AlHalwachi, Maryam Ghazi Alarayedh, and Amna Mohamed Buheiji who played an essential role in the data collection process related to this paper. Our thanks and appreciation goes to them for their hard, dedicated work. Wishing them all the best in the future. We would also like to extend our appreciation to Dr Simone Perna who dedicated time and effort to review and help us improve the manuscript.

