## Appendix ; Supplementary table for "The efficacy and safety of hydroxychloroquine in COVID19 patients : a multicenter national retrospective cohort"

### **Supplementary table: Disease severity clinical scale**

| **Score** | **Description** |
| --- | --- |
| 1 | Isolated and asymptomatic |
| 2 | Isolated and mildly asymptomatic |
| 3 | Admitted and on room air |
| 4 | Admitted and on oxygen support |
| 5 | Admitted and on NIV/HFNC |
| 6 | Admitted and on mechanical ventilation/ Extracorporeal membrane oxygenation |
| 7 | Death |

### **Appendix A: Collected Details from Patient Files**

Data collected in Questionnaire

| 1- **Demographic Data** | 2- **Comorbidities** | 3- **Symptoms on Admission** | 4- **Medication Taken** |
| --- | --- | --- | --- |
| Name | Sickle Cell Disease | Fever (>38C) | Hydroxychloroquine |
| Age | G6PD Deficiency | Cough | Azithromycin |
| CPR (ID Number) | Diabetes Mellitus | Chest pain | Kaletra |
| Gender | Cardiovascular Disease | Shortness Of breath | Ribavirin |
| Nationality | Hypertension | Loss of Smell | Zinc |
| Admission Date | Asthma / COPD | Loss of taste | Steroids |
| Admission Location | Other Chronic Lung Disease | Diarrhoea | Convalescent Plasma Transfusion |
|  | Obesity (BMI >=30) | Nausea or Vomiting | Tocilizumab |
|  | CKD | Body pain |  |
|  | Smoker |  |  |
| 5- **Vitals on Admission** | 6- **Oxygenation requirement on admission** | 7- **Chest X ray Finding on Admission** | 8- **Laboratory data on admission** |
| 9- **Clinical Status on Admission** | 10- **Clinical Outcome: NIV, Intubation, QT prolongation and Arrythmia.** | **11-Outcome: Death, Discharge** | **12- Adverse events** |

### **Appendix B: Propensity Score Details**

#### Propensity Score Regression Model

Creating the propensity score

**Logistic regression for receiving the treatment drug**

| Hydroxychloroquine | Coef. | | St.Err. | t-value | | p-value | [95% Conf | | Interval] | Sig |
| --- | --- | --- | --- | --- | --- | --- | --- | --- | --- | --- |
| Age | 1.048 | | 0.006 | 8.12 | | 0.000 | 1.036 | | 1.060 | *** |
| Male | 0.717 | | 0.101 | -2.36 | | 0.018 | 0.543 | | 0.945 | ** |
| SPO2:FIO2 Ratio | 0.998 | | 0.001 | -1.45 | | 0.148 | 0.996 | | 1.001 |  |
| Sickler Cell Disease | 0.464 | | 0.263 | -1.35 | | 0.176 | 0.153 | | 1.410 |  |
| G6PD | 0.282 | | 0.074 | -4.85 | | 0.000 | 0.169 | | 0.470 | *** |
| Diabetes Meletus | 1.074 | | 0.182 | 0.42 | | 0.672 | 0.771 | | 1.496 |  |
| Cardiovascular Disease | 0.760 | | 0.174 | -1.20 | | 0.231 | 0.485 | | 1.191 |  |
| Hypertension | 0.828 | | 0.150 | -1.04 | | 0.298 | 0.581 | | 1.181 |  |
| Asthma | 1.193 | | 0.403 | 0.52 | | 0.601 | 0.616 | | 2.313 |  |
| Chronic Obstructive Pulmonary Disease | 1.624 | | 1.792 | 0.44 | | 0.660 | 0.187 | | 14.115 |  |
| Obesity | 1.593 | | 0.604 | 1.23 | | 0.219 | 0.758 | | 3.351 |  |
| Chronic Kidney Disease | 1.619 | | 0.592 | 1.32 | | 0.187 | 0.791 | | 3.314 |  |
| Other Chronic Lung Disease | 0.834 | | 0.783 | -0.19 | | 0.847 | 0.132 | | 5.255 |  |
| Symptoms | 1.124 | | 0.225 | 0.58 | | 0.560 | 0.759 | | 1.664 |  |
| Fever | 1.528 | | 0.269 | 2.41 | | 0.016 | 1.083 | | 2.157 | ** |
| Cough | 1.534 | | 0.269 | 2.44 | | 0.015 | 1.088 | | 2.163 | ** |
| Chest Pain | 1.798 | | 0.418 | 2.52 | | 0.012 | 1.140 | | 2.837 | ** |
| Azithromycin | 10.727 | | 1.764 | 14.43 | | 0.000 | 7.772 | | 14.805 | *** |
| Kaletra | 2.076 | | 0.797 | 1.90 | | 0.057 | 0.978 | | 4.406 | * |
| Ribavirin | 0.066 | | 0.028 | -6.44 | | 0.000 | 0.029 | | 0.151 | *** |
| Constant | 0.044 | | 0.026 | -5.27 | | 0.000 | 0.014 | | 0.141 | *** |
| Mean dependent var | | 0.280 | | | SD dependent var | | | 0.449 | |  |
| Pseudo r-squared | | 0.258 | | | Number of obs | | | 1570.000 | |  |
| Chi-square | | 480.498 | | | Prob > chi2 | | | 0.000 | |  |
| Akaike crit. (AIC) | | 1424.131 | | | Bayesian crit. (BIC) | | | 1536.666 | |  |
|  | |  | | | area under ROC curve | | | 0.8325 | |  |
| *** p<0.01, ** p<0.05, * p<0.1 | | | | | | | | | |  |

#### **Histograms of Propensity Score Distribution in The Unmatched Sample**

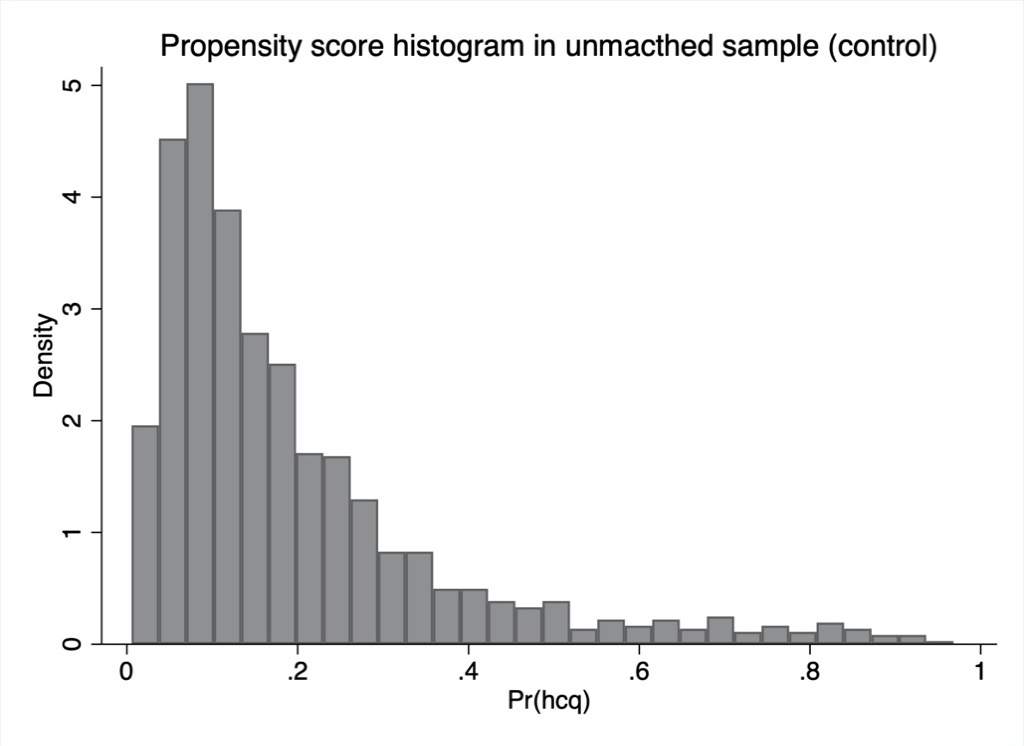

Figure 1a : propensity score histogram in the unmatched sample - control

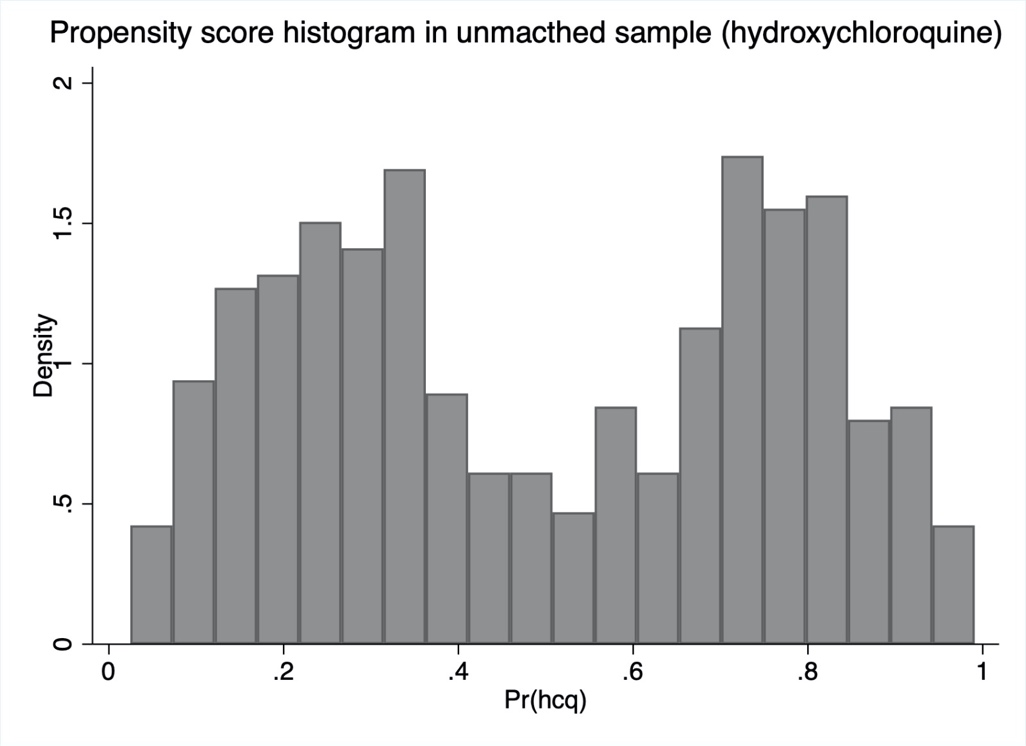

Figure 1b: propensity score histogram in the unmatched sample - HCQ

#### **Histograms of Propensity Score Distribution in The Matched Sample**

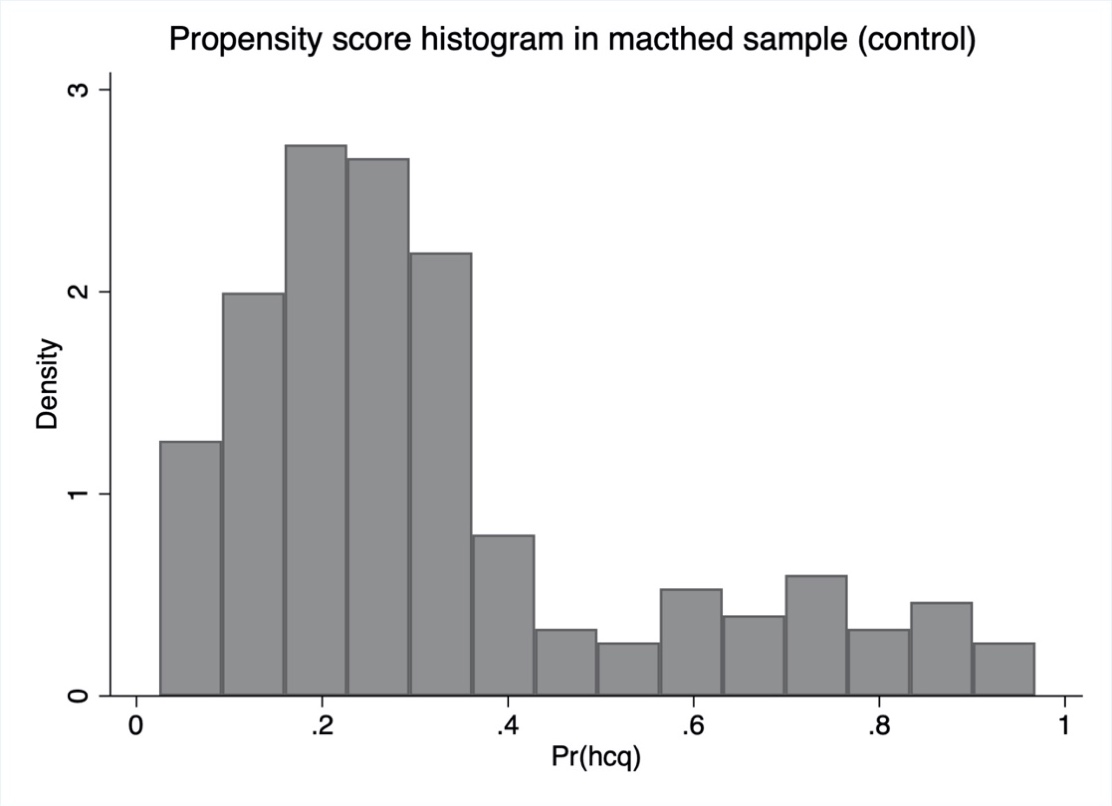

Figure 2a: propensity score histogram in the matched sample - control

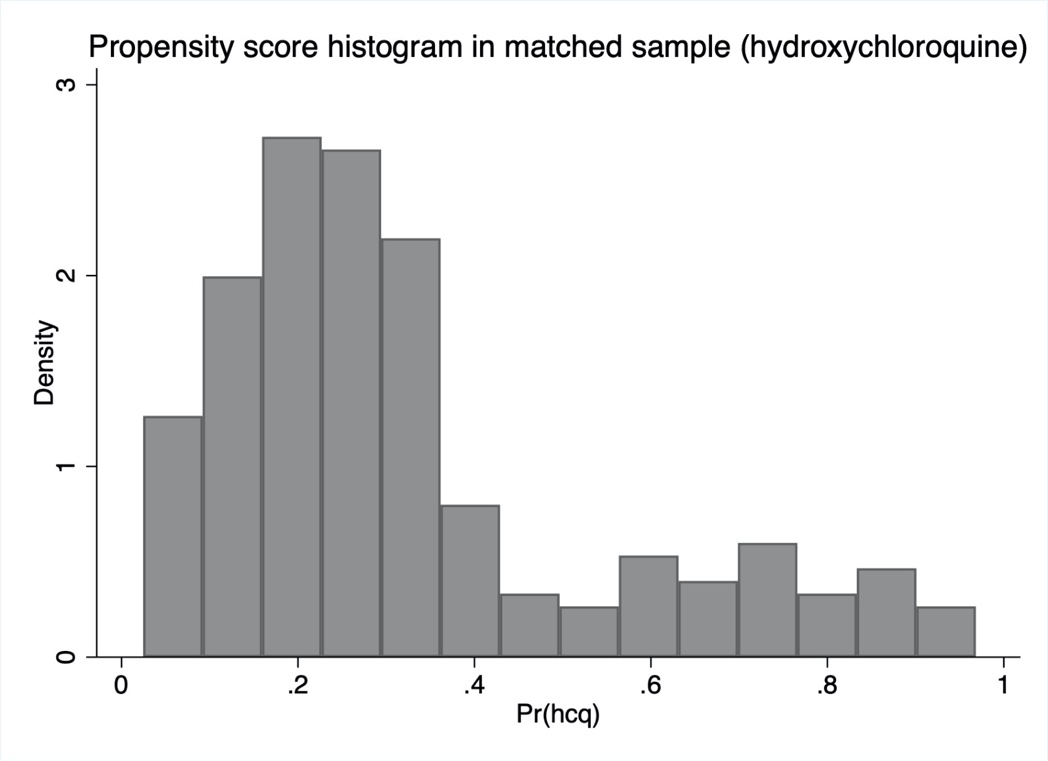

Figure 2b: propensity score histogram in the matched sample - control

### **Appendix C: Regression Models**

#### **Regression: Crude Analysis**

**Logistic regression**

| Primary composite outcome | Coef. | | St.Err. | t-value | | p-value | [95% Conf | | Interval] | Sig |
| --- | --- | --- | --- | --- | --- | --- | --- | --- | --- | --- |
| Hydroxychloroquine | 1.425 | | 0.371 | 1.36 | | 0.173 | 0.856 | | 2.374 |  |
| Constant | 0.040 | | 0.006 | -20.86 | | 0.000 | 0.030 | | 0.055 | *** |
| Mean dependent var | | 0.043 | | | SD dependent var | | | 0.204 | |  |
| Pseudo r-squared | | 0.003 | | | Number of obs | | | 1571.000 | |  |
| Chi-square | | 1.790 | | | Prob > chi2 | | | 0.181 | |  |
| Akaike crit. (AIC) | | 562.257 | | | Bayesian crit. (BIC) | | | 572.976 | |  |
|  | |  | | | area under ROC curve | | | 0.5381 | |  |
| *** p<0.01, ** p<0.05, * p<0.1 | | | | | | | | | |  |

#### **Regression: Multivariate Model Analysis**

**Logistic regression**

| Primary composite outcome | Coef. | | St.Err. | t-value | | p-value | [95% Conf | | Interval] | Sig |
| --- | --- | --- | --- | --- | --- | --- | --- | --- | --- | --- |
| Hydroxychloroquine | 1.646 | | 0.591 | 1.39 | | 0.166 | 0.814 | | 3.328 |  |
| Age | 1.016 | | 0.012 | 1.32 | | 0.187 | 0.992 | | 1.041 |  |
| Male | 1.064 | | 0.345 | 0.19 | | 0.849 | 0.563 | | 2.009 |  |
| SpO2:FiO2 Ratio | 0.992 | | 0.001 | -5.81 | | 0.000 | 0.990 | | 0.995 | *** |
| Comorbidities | 1.312 | | 0.142 | 2.51 | | 0.012 | 1.061 | | 1.621 | ** |
| Steroids | 6.714 | | 2.138 | 5.98 | | 0.000 | 3.597 | | 12.531 | *** |
| Ribavirin | 3.638 | | 1.331 | 3.53 | | 0.000 | 1.776 | | 7.454 | *** |
| Azithromycin | 2.410 | | 0.780 | 2.72 | | 0.007 | 1.278 | | 4.545 | *** |
| Constant | 0.030 | | 0.028 | -3.79 | | 0.000 | 0.005 | | 0.183 | *** |
| Mean dependent var | | 0.043 | | | SD dependent var | | | 0.202 | |  |
| Pseudo r-squared | | 0.402 | | | Number of obs | | | 1570.000 | |  |
| Chi-square | | 222.563 | | | Prob > chi2 | | | 0.000 | |  |
| Akaike crit. (AIC) | | 349.191 | | | Bayesian crit. (BIC) | | | 397.421 | |  |
|  | |  | | | area under ROC curve | | | 0.9366 | |  |
| *** p<0.01, ** p<0.05, * p<0.1 | | | | | | | | | |  |

#### **Regression: Propensity Score Adjustment Analysis** **Logistic regression**

| Primary composite outcome | Coef. | | St.Err. | t-value | | p-value | [95% Conf | | Interval] | Sig |
| --- | --- | --- | --- | --- | --- | --- | --- | --- | --- | --- |
| Hydroxychloroquine | 0.875 | | 0.281 | -0.42 | | 0.677 | 0.467 | | 1.641 |  |
| Propensity Score | 4.759 | | 2.536 | 2.93 | | 0.003 | 1.674 | | 13.525 | *** |
| Constant | 0.028 | | 0.006 | -17.41 | | 0.000 | 0.019 | | 0.042 | *** |
| Mean dependent var | | 0.043 | | | SD dependent var | | | 0.202 | |  |
| Pseudo r-squared | | 0.018 | | | Number of obs | | | 1570.000 | |  |
| Chi-square | | 10.151 | | | Prob > chi2 | | | 0.006 | |  |
| Akaike crit. (AIC) | | 549.602 | | | Bayesian crit. (BIC) | | | 565.679 | |  |
|  | |  | | | area under ROC curve | | | 0.6211 | |  |
| *** p<0.01, ** p<0.05, * p<0.1 | | | | | | | | | |  |

#### **Regression: Propensity Score Matched Analysis**

**Treatment-effects estimation**

| Primary Composite Outcome | Coef. | | St.Err. | t-value | | p-value | [95% Conf | | Interval] | Sig |
| --- | --- | --- | --- | --- | --- | --- | --- | --- | --- | --- |
| Hydroxychloroquine | -0.004 | | 0.012 | -0.36 | | 0.718 | -0.027 | | 0.019 |  |
| Mean dependent var | | 0.042 | | | SD dependent var | | | 0.200 | |  |
| *** p<0.01, ** p<0.05, * p<0.1 | | | | | | | | | |  |

#### **Regression: Oxygen Effect Modification Analysis**

**Logistic regression (with oxygen)**

| Primary composite outcome | Coef. | | St.Err. | t-value | | p-value | [95% Conf | | Interval] | Sig |
| --- | --- | --- | --- | --- | --- | --- | --- | --- | --- | --- |
| Hydroxychloroquine | 1.090 | | 0.577 | 0.16 | | 0.871 | 0.386 | | 3.078 |  |
| Age | 1.044 | | 0.018 | 2.45 | | 0.014 | 1.009 | | 1.080 | ** |
| Male | 1.080 | | 0.508 | 0.16 | | 0.870 | 0.430 | | 2.713 |  |
| SPO2:FIO2 Ratio | 0.989 | | 0.003 | -3.83 | | 0.000 | 0.984 | | 0.995 | *** |
| Comorbidities | 1.193 | | 0.172 | 1.23 | | 0.220 | 0.900 | | 1.583 |  |
| Steroids | 3.323 | | 1.446 | 2.76 | | 0.006 | 1.416 | | 7.799 | *** |
| Ribavirin | 1.066 | | 0.557 | 0.12 | | 0.902 | 0.383 | | 2.970 |  |
| Azithromycin | 2.299 | | 1.049 | 1.82 | | 0.068 | 0.940 | | 5.621 | * |
| Constant | 0.083 | | 0.112 | -1.85 | | 0.065 | 0.006 | | 1.162 | * |
| Mean dependent var | | 0.247 | | | SD dependent var | | | 0.433 | |  |
| Pseudo r-squared | | 0.249 | | | Number of obs | | | 178.000 | |  |
| Chi-square | | 49.561 | | | Prob > chi2 | | | 0.000 | |  |
| Akaike crit. (AIC) | | 167.524 | | | Bayesian crit. (BIC) | | | 196.161 | |  |
|  | |  | | | area under ROC curve | | | 0.8155 | |  |
| *** p<0.01, ** p<0.05, * p<0.1 | | | | | | | | | |  |

**Logistic regression: (without oxygen on baseline)**

| Primary composite outcome | Coef. | | St.Err. | t-value | | p-value | [95% Conf | | Interval] | Sig |
| --- | --- | --- | --- | --- | --- | --- | --- | --- | --- | --- |
| Hydroxychloroquine | 2.793 | | 1.574 | 1.82 | | 0.068 | 0.925 | | 8.431 | * |
| Age | 0.982 | | 0.019 | -0.94 | | 0.347 | 0.945 | | 1.020 |  |
| Male | 1.078 | | 0.522 | 0.15 | | 0.877 | 0.418 | | 2.784 |  |
| SPO2:FIO2 Ratio | 1.007 | | 0.008 | 0.90 | | 0.367 | 0.992 | | 1.022 |  |
| Comorbidities | 1.533 | | 0.259 | 2.53 | | 0.011 | 1.101 | | 2.135 | ** |
| Steroids | 14.952 | | 7.158 | 5.65 | | 0.000 | 5.850 | | 38.213 | *** |
| Ribavirin | 12.240 | | 6.328 | 4.84 | | 0.000 | 4.444 | | 33.717 | *** |
| Azithromycin | 2.439 | | 1.214 | 1.79 | | 0.073 | 0.920 | | 6.470 | * |
| Constant | 0.000 | | 0.000 | -2.66 | | 0.008 | 0.000 | | 0.071 | *** |
| Mean dependent var | | 0.017 | | | SD dependent var | | | 0.128 | |  |
| Pseudo r-squared | | 0.327 | | | Number of obs | | | 1392.000 | |  |
| Chi-square | | 76.621 | | | Prob > chi2 | | | 0.000 | |  |
| Akaike crit. (AIC) | | 175.735 | | | Bayesian crit. (BIC) | | | 222.882 | |  |
|  | |  | | | area under ROC curve | | | 0.9187 | |  |
| *** p<0.01, ** p<0.05, * p<0.1 | | | | | | | | | |  |

#### **Regression: Azithromycin Effect Modification Analysis**

**Logistic regression (with Azithromycin)**

| outcome | Coef. | | St.Err. | t-value | | p-value | [95% Conf | | Interval] | Sig |
| --- | --- | --- | --- | --- | --- | --- | --- | --- | --- | --- |
| Hydroxychloroquine | 2.706 | | 1.636 | 1.65 | | 0.100 | 0.827 | | 8.849 |  |
| Age | 1.022 | | 0.021 | 1.05 | | 0.293 | 0.981 | | 1.065 |  |
| Male | 1.675 | | 0.886 | 0.97 | | 0.329 | 0.594 | | 4.724 |  |
| SPO2:FIO2 Ratio | 0.992 | | 0.002 | -3.96 | | 0.000 | 0.988 | | 0.996 | *** |
| Comorbidities | 1.104 | | 0.179 | 0.61 | | 0.541 | 0.803 | | 1.518 |  |
| Steroids | 12.405 | | 6.308 | 4.95 | | 0.000 | 4.578 | | 33.609 | *** |
| Ribavirin | 5.086 | | 2.720 | 3.04 | | 0.002 | 1.783 | | 14.508 | *** |
| Azithromycin | 1.000 | | . | . | | . | . | | . |  |
| Constant | 0.042 | | 0.061 | -2.19 | | 0.029 | 0.002 | | 0.720 | ** |
| Mean dependent var | | 0.093 | | | SD dependent var | | | 0.291 | |  |
| Pseudo r-squared | | 0.407 | | | Number of obs | | | 386.000 | |  |
| Chi-square | | 97.413 | | | Prob > chi2 | | | 0.000 | |  |
| Akaike crit. (AIC) | | 157.927 | | | Bayesian crit. (BIC) | | | 189.574 | |  |
|  | |  | | | area under ROC curve | | | 0.9301 | |  |
| *** p<0.01, ** p<0.05, * p<0.1 | | | | | | | | | |  |

**Logistic regression (without Azithromycin)**

| outcome | Coef. | | St.Err. | t-value | | p-value | [95% Conf | | Interval] | Sig |
| --- | --- | --- | --- | --- | --- | --- | --- | --- | --- | --- |
| Hydroxychloroquine | 1.283 | | 0.703 | 0.46 | | 0.649 | 0.438 | | 3.753 |  |
| Age | 1.015 | | 0.016 | 0.93 | | 0.353 | 0.984 | | 1.046 |  |
| Male | 0.705 | | 0.308 | -0.80 | | 0.423 | 0.300 | | 1.659 |  |
| SPO2:FIO2 Ratio | 0.991 | | 0.002 | -4.47 | | 0.000 | 0.987 | | 0.995 | *** |
| Comorbidities | 1.591 | | 0.244 | 3.02 | | 0.003 | 1.177 | | 2.150 | *** |
| Steroids | 4.880 | | 2.298 | 3.37 | | 0.001 | 1.939 | | 12.283 | *** |
| Ribavirin | 2.525 | | 1.358 | 1.72 | | 0.085 | 0.880 | | 7.244 | * |
| Azithromycin | 1.000 | | . | . | | . | . | | . |  |
| Constant | 0.041 | | 0.050 | -2.60 | | 0.009 | 0.004 | | 0.455 | *** |
| Mean dependent var | | 0.026 | | | SD dependent var | | | 0.160 | |  |
| Pseudo r-squared | | 0.368 | | | Number of obs | | | 1184.000 | |  |
| Chi-square | | 105.677 | | | Prob > chi2 | | | 0.000 | |  |
| Akaike crit. (AIC) | | 197.349 | | | Bayesian crit. (BIC) | | | 237.963 | |  |
|  | |  | | | area under ROC curve | | | 0.9369 | |  |
| *** p<0.01, ** p<0.05, * p<0.1 | | | | | | | | | |  |

**Regression: Multivariate Model for Adverse Events Analysis**

**Logistic regression (Hypoglycemia):**

| Hypoglycemia | Coef. | | St.Err. | t-value | | p-value | [95% Conf | | Interval] | Sig |
| --- | --- | --- | --- | --- | --- | --- | --- | --- | --- | --- |
| Hydroxychloroquine | 10.937 | | 10.318 | 2.54 | | 0.011 | 1.722 | | 69.486 | ** |
| Age | 1.062 | | 0.029 | 2.23 | | 0.026 | 1.007 | | 1.120 | ** |
| Male | 11.122 | | 12.491 | 2.15 | | 0.032 | 1.231 | | 100.505 | ** |
| SPO2:FIO2 Ratio | 1.008 | | 0.006 | 1.25 | | 0.213 | 0.995 | | 1.021 |  |
| Comorbidities | 1.311 | | 0.296 | 1.20 | | 0.230 | 0.842 | | 2.039 |  |
| Steroids | 1.018 | | 1.143 | 0.02 | | 0.988 | 0.113 | | 9.192 |  |
| Ribavirin | 9.168 | | 7.400 | 2.75 | | 0.006 | 1.885 | | 44.601 | *** |
| Azithromycin | 1.078 | | 0.840 | 0.10 | | 0.924 | 0.234 | | 4.966 |  |
| Constant | 0.000 | | 0.000 | -4.40 | | 0.000 | 0.000 | | 0.000 | *** |
| Mean dependent var | | 0.006 | | | SD dependent var | | | 0.076 | |  |
| Pseudo r-squared | | 0.307 | | | Number of obs | | | 1570.000 | |  |
| Chi-square | | 34.079 | | | Prob > chi2 | | | 0.000 | |  |
| Akaike crit. (AIC) | | 94.778 | | | Bayesian crit. (BIC) | | | 143.007 | |  |
|  | |  | | | area under ROC curve | | | 0.9178 | |  |
| *** p<0.01, ** p<0.05, * p<0.1 | | | | | | | | | |  |

**Logistic regression (Diarrhea):**

| Diarrhea | Coef. | | St.Err. | t-value | | p-value | [95% Conf | | Interval] | Sig |
| --- | --- | --- | --- | --- | --- | --- | --- | --- | --- | --- |
| Hydroxychloroquine | 2.807 | | 0.973 | 2.98 | | 0.003 | 1.423 | | 5.538 | *** |
| Age | 0.996 | | 0.012 | -0.33 | | 0.739 | 0.974 | | 1.019 |  |
| Male | 0.822 | | 0.251 | -0.64 | | 0.521 | 0.453 | | 1.494 |  |
| SPO2:FIO2 Ratio | 0.999 | | 0.002 | -0.65 | | 0.515 | 0.996 | | 1.002 |  |
| Comorbidities | 1.121 | | 0.113 | 1.13 | | 0.257 | 0.920 | | 1.367 |  |
| Steroids | 1.588 | | 0.629 | 1.17 | | 0.243 | 0.730 | | 3.452 |  |
| Ribavirin | 3.181 | | 1.176 | 3.13 | | 0.002 | 1.541 | | 6.567 | *** |
| Azithromycin | 2.202 | | 0.724 | 2.40 | | 0.016 | 1.156 | | 4.195 | ** |
| Constant | 0.016 | | 0.017 | -3.96 | | 0.000 | 0.002 | | 0.124 | *** |
| Mean dependent var | | 0.032 | | | SD dependent var | | | 0.176 | |  |
| Pseudo r-squared | | 0.106 | | | Number of obs | | | 1570.000 | |  |
| Chi-square | | 46.971 | | | Prob > chi2 | | | 0.000 | |  |
| Akaike crit. (AIC) | | 414.100 | | | Bayesian crit. (BIC) | | | 462.330 | |  |
|  | |  | | | area under ROC curve | | | 0.7847 | |  |
| *** p<0.01, ** p<0.05, * p<0.1 | | | | | | | | | |  |

**Logistic regression (QT Prolongation):**

| QT Prolongation | Coef. | | St.Err. | t-value | | p-value | [95% Conf | | Interval] | Sig |
| --- | --- | --- | --- | --- | --- | --- | --- | --- | --- | --- |
| Hydroxychloroquine | 1.925 | | 0.692 | 1.82 | | 0.069 | 0.951 | | 3.896 | * |
| Age | 1.032 | | 0.012 | 2.70 | | 0.007 | 1.009 | | 1.055 | *** |
| Male | 0.369 | | 0.118 | -3.12 | | 0.002 | 0.198 | | 0.690 | *** |
| SPO2:FIO2 Ratio | 0.999 | | 0.002 | -0.37 | | 0.715 | 0.996 | | 1.002 |  |
| Comorbidities | 0.993 | | 0.100 | -0.07 | | 0.942 | 0.815 | | 1.208 |  |
| Steroids | 1.769 | | 0.650 | 1.55 | | 0.121 | 0.860 | | 3.636 |  |
| Ribavirin | 14.484 | | 5.080 | 7.62 | | 0.000 | 7.284 | | 28.803 | *** |
| Azithromycin | 1.046 | | 0.356 | 0.13 | | 0.896 | 0.537 | | 2.037 |  |
| Constant | 0.004 | | 0.004 | -5.66 | | 0.000 | 0.001 | | 0.026 | *** |
| Mean dependent var | | 0.035 | | | SD dependent var | | | 0.184 | |  |
| Pseudo r-squared | | 0.264 | | | Number of obs | | | 1570.000 | |  |
| Chi-square | | 125.971 | | | Prob > chi2 | | | 0.000 | |  |
| Akaike crit. (AIC) | | 368.744 | | | Bayesian crit. (BIC) | | | 416.973 | |  |
|  | |  | | | area under ROC curve | | | 0.8736 | |  |
| *** p<0.01, ** p<0.05, * p<0.1 | | | | | | | | | |  |

**Logistic regression (Arrythmia):**

| Arrythmia | Coef. | | | St.Err. | t-value | | p-value | [95% Conf | | Interval] | Sig |
| --- | --- | --- | --- | --- | --- | --- | --- | --- | --- | --- | --- |
| Hydroxychloroquine | | 1.062 | | 0.356 | 0.18 | | 0.857 | 0.551 | | 2.048 |  |
| Age | 1.025 | | | 0.011 | 2.34 | | 0.019 | 1.004 | | 1.046 | ** |
| Male | 0.942 | | | 0.272 | -0.21 | | 0.836 | 0.534 | | 1.660 |  |
| SPO2:FIO2 Ratio | 0.998 | | | 0.001 | -1.58 | | 0.115 | 0.995 | | 1.001 |  |
| Comorbidities | 1.013 | | | 0.096 | 0.14 | | 0.889 | 0.842 | | 1.219 |  |
| Steroids | 2.793 | | | 0.933 | 3.07 | | 0.002 | 1.451 | | 5.376 | *** |
| Ribavirin | 2.476 | | | 0.864 | 2.60 | | 0.009 | 1.250 | | 4.905 | *** |
| Azithromycin | 1.345 | | | 0.425 | 0.94 | | 0.348 | 0.725 | | 2.497 |  |
| Constant | 0.016 | | | 0.015 | -4.50 | | 0.000 | 0.003 | | 0.098 | *** |
| Mean dependent var | | | 0.036 | | | SD dependent var | | | 0.187 | |  |
| Pseudo r-squared | | | 0.111 | | | Number of obs | | | 1570.000 | |  |
| Chi-square | | | 54.571 | | | Prob > chi2 | | | 0.000 | |  |
| Akaike crit. (AIC) | | | 453.333 | | | Bayesian crit. (BIC) | | | 501.563 | |  |
|  | | |  | | | area under ROC curve | | | 0.7381 | |  |
| *** p<0.01, ** p<0.05, * p<0.1 | | | | | | | | | | |  |

#### **Regression: Secondary Outcome Length of Stay**

Secondary Outcome Length of stay

**Linear regression: crude Analysis**

| Length of stay | Coef. | | St.Err. | t-value | | p-value | [95% Conf | | Interval] | Sig |
| --- | --- | --- | --- | --- | --- | --- | --- | --- | --- | --- |
| Hydroxychloroquine | 1.793 | | 0.311 | 5.76 | | 0.000 | 1.183 | | 2.403 | *** |
| Constant | 9.492 | | 0.165 | 57.54 | | 0.000 | 9.168 | | 9.816 | *** |
| Mean dependent var | | 9.996 | | | SD dependent var | | | 5.539 | |  |
| R-squared | | 0.021 | | | Number of obs | | | 1536.000 | |  |
| F-test | | 33.214 | | | Prob > F | | | 0.000 | |  |
| Akaike crit. (AIC) | | 9587.546 | | | Bayesian crit. (BIC) | | | 9598.219 | |  |
| *** p<0.01, ** p<0.05, * p<0.1 | | | | | | | | | |  |

**Linear regression: multivariate model**

| Length of stay | Coef. | | St.Err. | t-value | | p-value | [95% Conf | | Interval] | Sig |
| --- | --- | --- | --- | --- | --- | --- | --- | --- | --- | --- |
| Hydroxychloroquine | 0.565 | | 0.316 | 1.79 | | 0.074 | -0.055 | | 1.185 | * |
| Age | 0.016 | | 0.009 | 1.79 | | 0.074 | -0.002 | | 0.034 | * |
| Male | 0.024 | | 0.247 | 0.10 | | 0.921 | -0.460 | | 0.509 |  |
| SPO2:FIO2 Ratio | 0.001 | | 0.002 | 0.41 | | 0.681 | -0.004 | | 0.005 |  |
| Comorbidities | 0.097 | | 0.083 | 1.16 | | 0.245 | -0.066 | | 0.260 |  |
| Steroids | 2.631 | | 0.458 | 5.74 | | 0.000 | 1.732 | | 3.529 | *** |
| Ribavirin | 2.950 | | 0.415 | 7.11 | | 0.000 | 2.136 | | 3.764 | *** |
| Azithromycin | 2.059 | | 0.324 | 6.36 | | 0.000 | 1.423 | | 2.694 | *** |
| outcome | 12.028 | | 0.890 | 13.51 | | 0.000 | 10.281 | | 13.774 | *** |
| Constant | 6.936 | | 1.167 | 5.94 | | 0.000 | 4.646 | | 9.226 | *** |
| Mean dependent var | | 9.996 | | | SD dependent var | | | 5.539 | |  |
| R-squared | | 0.276 | | | Number of obs | | | 1536.000 | |  |
| F-test | | 64.766 | | | Prob > F | | | 0.000 | |  |
| Akaike crit. (AIC) | | 9139.528 | | | Bayesian crit. (BIC) | | | 9192.897 | |  |
| *** p<0.01, ** p<0.05, * p<0.1 | | | | | | | | | |  |
